# The ABCs of Subarachnoid Hemorrhage Blood Volume Measurement: A Simplified Quantitative Method Predicts Outcomes and Delayed Cerebral Ischemia

**DOI:** 10.1101/2023.09.05.23295090

**Authors:** Fabian Foettinger, Rohan Sharma, Saif Salman, Alexander D. Weston, Bradley J. Erickson, Thien Huynh, Rabih G. Tawk, William D. Freeman

## Abstract

**Background:** We sought to develop an easy-to-use quantifiable and more precise method to estimate total blood volume in patients with aneurysmal subarachnoid hemorrhage (SAH) and explore its volumetric extent on incidence of delayed cerebral ischemia (DCI) and poor outcomes by modified Rankin Scale by hospital discharge.

**Methods:** In this retrospective observational cohort study, we analyzed 277 patients with SAH admitted at our Comprehensive Stroke Center between 2012 and 2022. We derived a mathematical model (Model 1) by measuring SAH basal cisternal blood volume using a ABC/2-derived ellipsoid formula (A=width/thickness, B=length, C=vertical extension) on noncontrast computed tomography (NCCT) in 5 major SAH cisternal compartments. We compared Model 1 (M1) against a manual segmentation method (Model 2) on noncontrast head CT (NCCT). Data were analyzed using logistic regression analysis, *t* test, receiver operator characteristic curves (ROC), and area under curve (AUC) analysis.

**Results:** There was no significant difference in cisternal SAH volume (SAHV) analysis between the two models (p=.14). Average SAHV by M1 was 7.0 mL (95% CI, 5,89-8,09) and lower for good outcome at discharge and 16.6 mL (95% CI, 13.49-19.77) for poor outcome. Volumetric analysis showed that patients with DCI had higher SAHV with 10 mL as a cutoff value (J_ABC-SAHV-outcome_ =10.0855 mL and J_ABC-SAHV-DCI_ =10.117 mL).

**Conclusions:** A simplified ABC/2-derived method of SAH blood volume measurement is comparable to manual segmentation, and more importantly, can be performed in low-resource settings. Higher total SAH volumes by ABC/2-derived method were associated with worse outcomes and higher risk of DCI in this study population. A potential dose-response relationship of SAHV blood greater than 10mL appeared to predict worse outcomes and higher risk for DCI. A future larger prospective trial is being planned to validate these results.

**Subject codes:** [50] Cerebral Aneurysm, AVM, & Subarachnoid hemorrhage; [58] Computerized tomography and Magnetic Resonance Imaging

## Introduction

Subarachnoid hemorrhage (SAH) is a devastating subtype of hemorrhagic stroke that is associated with a high one-month mortality rate of 30% to 40% and one that requires a more individualized medicine approach as to why some SAH patients. Delayed cerebral ischemia (DCI), a frequently encountered secondary complication of SAH, has been demonstrated to be associated with compromised long-term outcomes, higher modified Rankin Scale (mRS) at discharge, cognitive impairment, and increased length of hospitalization, and substantially increases risks of morbidity and mortality.[1, 2] Recent trends propagate a multifactorial pathophysiology of DCI, including vasospasm, cortical spreading depolarization, micro-thrombosis, neuroinflammation, and cerebrovascular dysregulation that result in parenchymal damage.[3] As for prognostication of poor functional outcome and DCI, prior studies have found a strong association between radiologically quantified SAH volume, clot thickness, hemorrhagic persistence, and concomitant intraventricular hemorrhage (IVH).[4-6]

The Fisher Scale (FS) and the modified Fisher Scale (mFS) arguably stand as the most prevalent radiographic systems in common clinical utilization [7-9] However, due to the lack of robust metric definitions, poor discriminative strength and questionable inter-rater reliability, newer radiologic scaling systems that improve clinical prediction of outcomes and DCI would help address a major unmet patient need.[10, 11] Recent studies propose that manual segmentation of neuroimaging allows for accurate quantification of hemorrhagic volume in SAH and prediction of poor outcome and secondary complications. However, the time-and resource-consuming nature of these segmentation methods and their technical training requirements compromise their clinical applicability.[12, 13] Therefore, an alternative simplified, but clinically feasible quantitative method by applying mathematical principles used in other neurovascular imaging research[14, 15] was conceptualized and explored in this study.

## Methods

### Study population

We retrospectively reviewed the electronic health records of 277 patients with aneurysmal SAH seen at Mayo Clinic in Jacksonville, Florida, between January 5, 2012, and February 24, 2022. Demographics, medical history, clinical assessment, computed tomography (CT), and hospital course were analyzed. Inclusion criteria for the study required non-contrast CT (NCCT) head imaging to be done at admission within 24 hours of SAH ictus and aneurysmal cause of SAH to be diagnosed through vascular imaging such as CT or MR angiogram, or cerebral angiogram. Furthermore, NCCT data had to have imaging slices of 5 mm or less or at least equivalent reformatting to adequately measure volumetric data quantitatively. If DCI was suspected, vascular neuroimaging modalities such as computed tomography angiography, magnetic resonance imaging, magnetic resonance angiography or digital subtraction angiography were employed to establish a diagnosis and to definitively rule out ischemic stroke. Criteria for exclusion included the absence of NCCT imaging, no diagnosis of SAH or only associated disease (only intracerebral hemorrhage in the medical record, IVH or subdural hemorrhage without SAH) and traumatic SAH.

This single-center, retrospective study was approved by the Mayo Clinic Institutional Review Board (IRB) since it utilized pre-existing subjects within the Mayo Clinic firewall along with stored clinical, laboratory, and radiographic data.

### Outcome and DCI

Outcome was quantified via modified Rankin Scale (mRS) score at discharge and dichotomized into good outcome (mRS 0-3) and poor outcome (mRS 4-6). DCI was defined in accordance the consensus definition by Vergouwen et al as as the occurrence of focal neurological impairment or a decrease of at least two points on GCS, persisting for more than an hour that is not caused by other clinically assessable conditions as well as intra- or postoperative changes or complications.[16] If patients died within 72 hours after admission, they were excluded from analysis of DCI.

### Imaging Analysis

The extraction of patient data, as well as the analysis of imaging data, were conducted by the principal investigator. To reduce the risk of bias, each analytic segment of this study was performed separately from another.

Two methods of SAHV (SAH Volume) estimation were conceptualized: the first was a *s*implified *Q*uantitative *v*olumetric model (sQv or ABC/2-derived **SAHV**, Model 1 [M1]) derived by ABC/2 mathematical linear measurements in 3 dimensions. The second was a *m*anual *s*egmentation *m*odel (msQv) also called segment-SAHV or Model 2[M2]) method. For matters of comparability, both methods were applied to the same NCCT images of the enrolled patients. The analyzed images were stored either in Digital Imaging and Communications in Medicine (DICOM) format or Neuroimaging Informatics Technology Initiative (NIfTI) format, as appropriate. Imaging analysis was performed on a personal computer using medical imaging software “QReads” for determination of metric extensions and open-source segmentation software “RIL Contour” (Version 0.1.947 Mayo Digital Innovation Lab, Mayo Clinic, Jacksonville, United States of America) for manual segmentation.

### Model 1: ABC/2-derived SAHV Methodology

Subarachnoid blood volume (SAHV) was measured in the following 5 predefined anatomic regions: prepontine cistern, perimesencephalic cistern, suprasellar cistern, left and right Sylvian fissure, middle cerebral artery fissure, and anterior interhemispheric fissure. (**Figure 1**) In 3 measurements, A = a linear measurement in the long axis of the SAH blood, B = thickest portion in cross-section, and C = vertical dimensions on the number of CT slices. Similarly to the ABC/2-volume measurement tool [15] for intracerebral hemorrhage (ICH), cylindrical volumetrics can be estimated by calculating volume through application of ABC/2 formula in 3 dimensions (ie., A = width, B = length, C = vertical dimension) and assumes that the basal cisterns “star” shaped morphology can be compartmentalized into subsegmental ellipsoid volumes (Model 1). ABC/2 is a simplified derivation of the ellipsoid formula and has previously been proven to yield accurate volumetric estimations in ICH and subdural hematoma (SDH).[14, 15, 17] The first step in measuring SAH hemorrhage was obtaining each of the 3 dimensions for each of the 5 compartments. For example, the maximal thickness (A) and length (B) for each individual cistern. Then, the number of slices vertically was counted containing discernible SAH blood and subsequently multiplied by the slice thickness (mm) of the given CT-image modality to account for the vertical extension (C). These 3 variables were then inputted into ABC/2 (**Figure 1**) equation to obtain the simplified volumetric estimate for each of the 5 cisternal compartments and the sum of the 5 were then calculated for each NCCT. Due to the inherent morphologic limitations of the ventricular systems and complex hemorrhagic distribution patterns, we did not include IVH volumetrics in this study. We did obtained modified Graeb IVH volumes during NCCT measurements and did not attempt to derive ellipsoid volumetrics for these irregular and often S-shaped hemorrhages.

**Figure 1.**
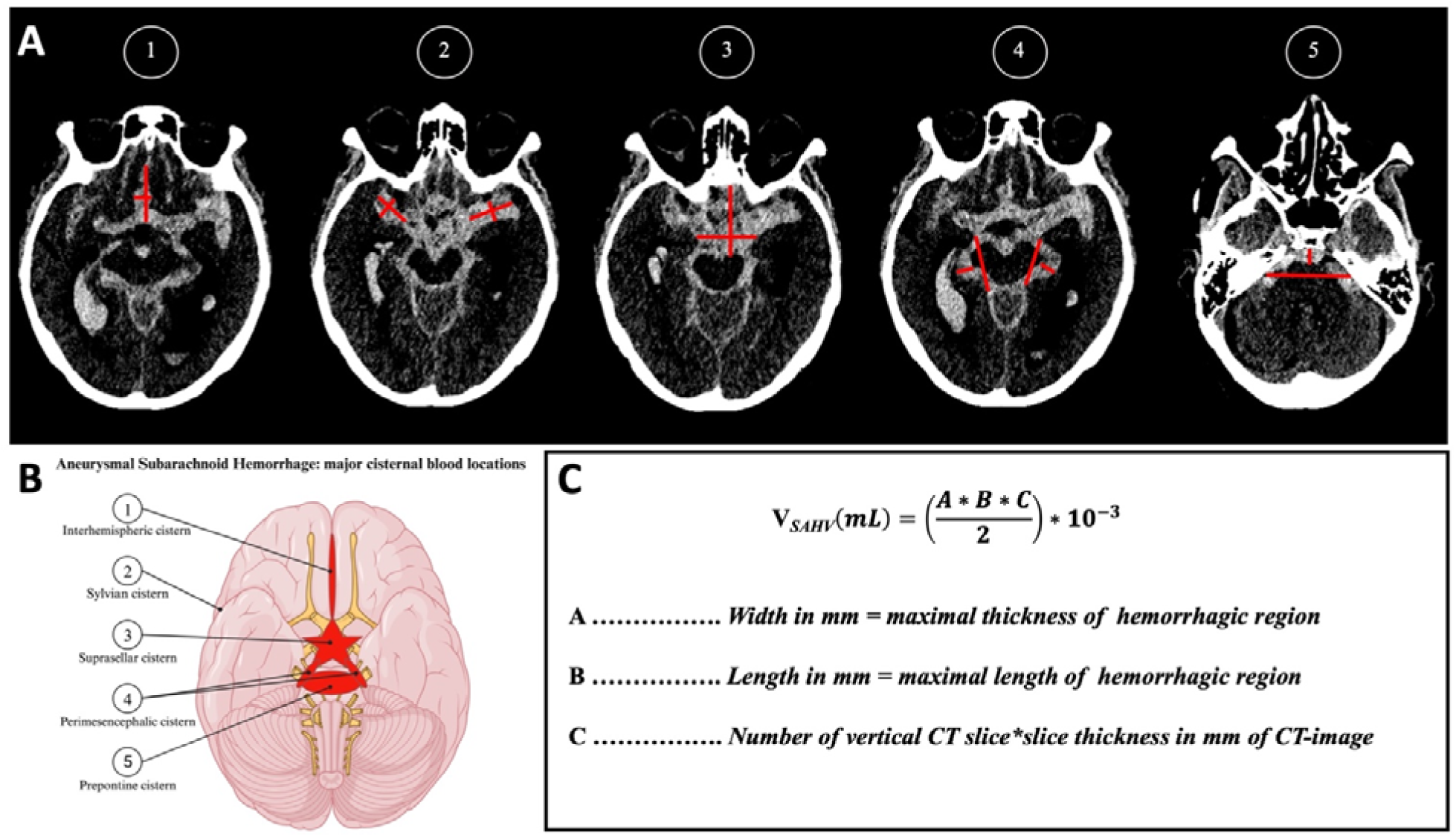
Volumetric Analysis Via ABC/2-derived SAHV. ***A,*** Multiple noncontrast CT slices of a patient with aneurysmal subarachnoid hemorrhage. There are visible SAH hyperdensities in multiple cisternal compartments, marked with lines to determine length and width of the respected hemorrhage in mm. Measurement of width, length, and vertical extension (number of NCCT slices with visible SAH) was done in consideration of cisternal anatomy. ***B*** Illustration of 5 SAHV compartments measured (Created with BioRender.com). ***C,*** These metric variables were then inputted into our Model 1 ABC/2-derived SAHV equation to measure the hemorrhagic volume in each neuroanatomic structure. Each volume was then summed to a cumulative total SAHV. SAHV indicates cisternal subarachnoid hemorrhage volume; NCCT, noncontrast computed tomography.

### Model 2: Segmentation SAHV Methodology

The same NCCT images were then separately analyzed by applying a manual segmentation approach using open-source RIL-Contour and Anaconda[18]. For Model 2 SAHV, a region of interest technique was applied, and SAHV-M2 manually contoured slice by slice. Voxels within these marked or *painted* areas were then translated into metric scales of mm and SAHV was calculated under consideration of the vertical extension and slice thickness of NCCT imaging. The same 5 predefined neuroanatomic compartments were measured using this Model 2 method and summated to calculate the total blood for SAHV-M2.

### Statistical Analysis

Patient characteristics were dichotomized based on their outcome at discharge and descriptively analyzed. Differences in categorical variables were determined via Fisher exact test, and differences in continuous variables were analyzed using paired/unpaired *t* test or Mann-Whitney U test, as appropriate. Descriptive statistics were used to depict volume estimations. To assess our SAHV models predictiveness of poor functional outcome and DCI multiple logistic regression analysis was performed with a classification cutoff of 0.5. Outcome in multiple logistic regression was dichotomized based on the patients’ mRS score at discharge. Differences in discriminative accuracy of SAHV and modified Fisher Scale were examined via receiver operator characteristic (ROC) curves and subsequent area under the curve (AUC) analysis. We also proposed specific outcome and DCI cutoff values for SAHV-M1 using Youden index (J = sensitivity + specificity – 1 or J = [true positives]/[true positives = false negatives] + [true negatives]/[true negative = false positives] − 1) by determining optimum cut points in ROC curve analysis. For statistical analysis, SPSS (IBM) and GraphPad (GraphPad Software) were used on a Windows-operated personal computer, and a *P* value of .05 was considered statistically significant.

## Results

### Patient Demographics

Among the initial 277 patients with SAH in our dataset, we excluded 72 due to traumatic SAH or non-aneurysmal SAH, lack of imaging quality and transferal to other hospitals, for a total study population of 205 patients with SAH. Patient characteristics are shown in **Table 1**. Median (range) Glasgow Coma Scale scores on admission were 15 (13-15) in the good outcome group and 7 (7-15) in the poor outcome group (p <.0001). Only 24 patients (11.7%) presented with concomitant IPH, with most being in the poor outcome group (20 [83.3%]). Forty-one patients (20.0%) had symptomatic (clinical) vasospasm (sVSP), and most vasospasm events occurred in the poor outcome group (27 [65.9%]). Higher modifed Graeb score of IVH predicted worse outcome (P < 0.001). DCI was more frequently encountered than symptomatic vasospasm (51 [24.4%]), and most patients with DCI (43 [84.3%]) were discharged with poor outcomes The predominant mFS grading observed in this study among all patients was mFS 4 (121 [59.0%]), followed by a mFS score 3 (48 [23.4%]).

**Table 1.** Baseline Patient Characteristics.

|  | Total<br>(N=205) | Dichotomized functional outcome at discharge |  |  |
| --- | --- | --- | --- | --- |
|  |  | Good outcome<br>(n=115) | Poor outcome<br>(n=90) | *P* value |
| Age, mean (SD), y | 56 (14) | 52 (12) | 61 (14) | <.0001<sup>a</sup> |
| Sex, No. (%), female | 133 (64.9) | 81 (70.4) | 53 (58.9) | .11<sup>b</sup> |
| Hypertension, No. (%) | 121 (59.0) | 69 (60.0) | 52 (57.8) | .78<sup>b</sup> |
| CVD, No. (%) | 25 (64.9) | 11 (9.6) | 14 (15.6) | .21<sup>b</sup> |
| Type 2 diabetes, No. (%) | 16 (7.8) | 10 (8.7) | 6 (6.7) | .79<sup>b</sup> |
| Smoking, No. (%) |  |  |  |  |
| Non-smoker | 76 (37.1) | 38 (33.0) | 38 (42.2) | .19<sup>b</sup> |
| Former smoker | 44 (21.5) | 30 (26.1) | 15 (16.7) | .13<sup>b</sup> |
| Active at SAH | 84 (41.0) | 46 (40.0) | 38 (42.2) | .78<sup>b</sup> |
| Alcohol use, No. (%) | 40 (19.5) | 22 (19.1) | 18 (20.0) | 1.00<sup>b</sup> |
| GCS score at admission, median (IQR) | 14 (7-15) | 15 (13-15) | 7 (4-15) | <.0001<sup>a</sup> |
| Hunt and Hess at admission, median (IQR) | 3 (2-4) | 2 (2-3) | 4 (3-5) | <.0001<sup>a</sup> |
| IPH, No. (%) | 24 (11.7) | 4 (3.5) | 20 (22.2) | <.0001<sup>b</sup> |
| Modified Fisher Scale score, No. (%) |  |  |  |  |
| 1 | 19 (9.3) | 18 (15.7) | 1 (1.1) |  |
| 2 | 15 (7.3) | 14 (12.2) | 1 (1.1) |  |
| 3 | 48 (23.4) | 33 (28.7) | 15 (16.7) |  |
| 4 | 121 (59.0) | 48 (41.7) | 74 (82.2) |  |
| Modified Graeb Scale score, median (IQR) | 6 (0-15) | 2 (0-8) | 13 (6-20) | <.0001<sup>a</sup> |
| Symptomatic vasospasm, No. (%) | 41 (20.0) | 14 (12.2) | 27 (30.0) | <.003<sup>b</sup> |
| DCI, No. (%) | 51 (24.4) | 7 (6.1) | 43 (47.8) | .001<sup>b</sup> |
| Abbreviations: CVD, cardiovascular disease; DCI, delayed cerebral ischemia; GCS, Glasgow Coma Scale; IPH, intraparenchymal hemorrhage; SAH, subarachnoid hemorrhage. |  |  |  |  |
| <sup>a</sup> Significance testing: Mann-Whitney-U test |  |  |  |  |
| <sup>b</sup> Significance testing: Fisher exact test |  |  |  |  |

### Volumetric Analysis

ABC/2-derived SAHV in all 205 patients were estimated using our Model 1 methodology. Mean SAHV was 11.3 mL (95% CI, 9.58-12.85) and the respective range was 125 mL, with a minimum of 0 mL (when no blood was present at the level of the cistern) and a maximum of 125 mL. Comparison of Model 2 (M2) with manually segmented CT images is shown in **Figure 2**. The mean ABC/2-derived SAHV measured by Model 1 was not significantly different than manually segmented SAHV-M2 (*P*=.14 and *P*=.21, respectively, **Figure 2**).

**Figure 2.**
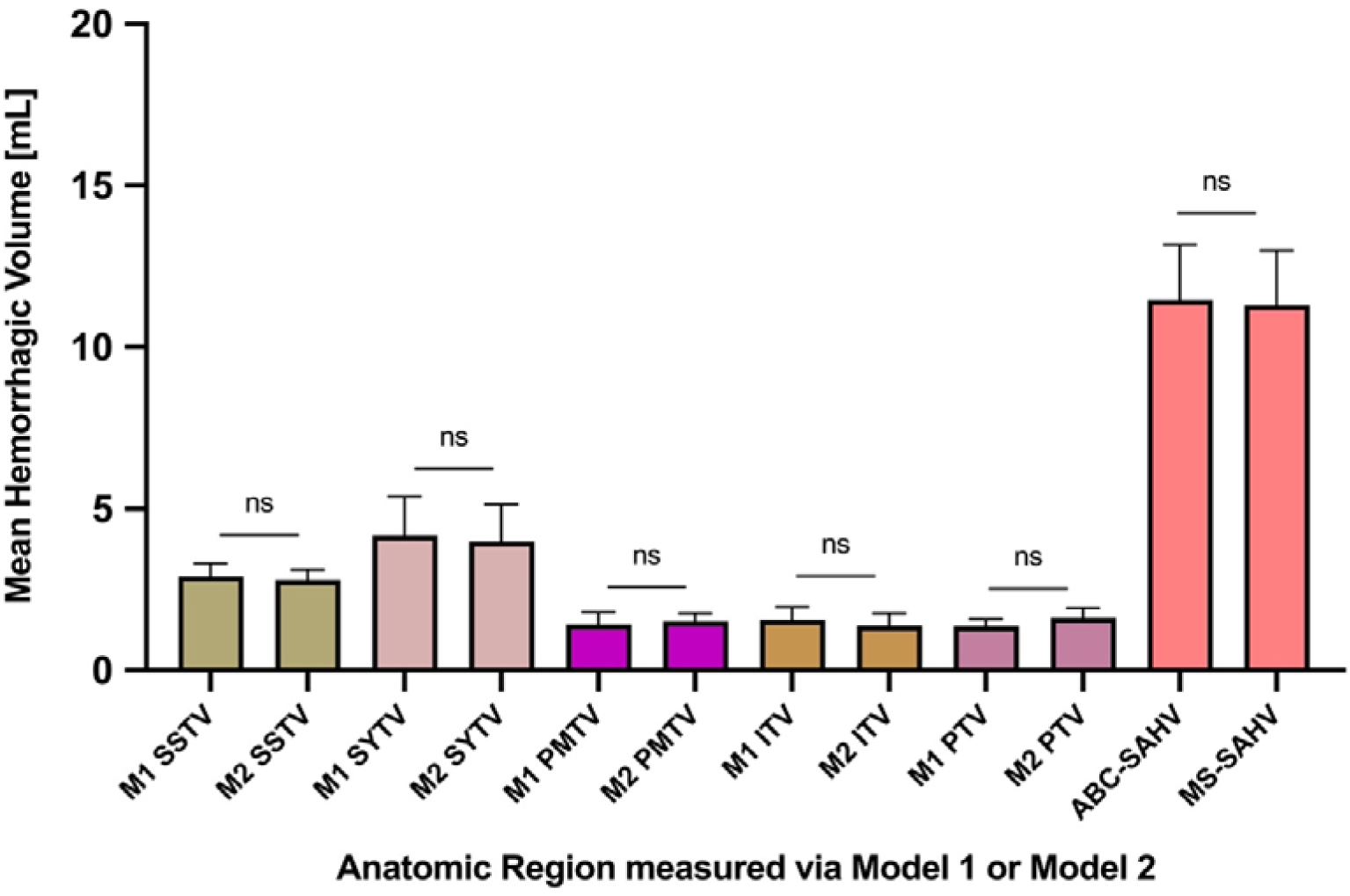
Comparison of hemorrhagic volumes via volumetric estimation using ABC/2 derived SAHV (M1) vs segment-SAHV (M2). Average volume per cistern depending on different methods applied. 5 SAH spaces or compartments: SSTV = suprasellar total volume; SYTV = Sylvian total volume; PMTV = perimesencephalic total volume; ITV = interhemispheric total volume; PTV = pontine total volume; significance testing: unpaired t-test.

### Association of Volume with Outcome and DCI

The volumetric relationship between predictor variables and dichotomized target variables is shown in **Figure 3**. For Model 1, the mean SAHV at discharge for the good outcome group was 7.0 mL (95% CI, 5.89-8.09), whereas the poor outcome group had significantly higher volumes with a mean SAHV of 16.6 mL (95% CI, 13.49-19.77). These differences in SAH volume are statistically significant (*P*<.05). Volumetric analysis for patients with DCI showed significantly higher SAH volumes of compared to patients without DCI for both SAHV-M1 (*P*<.05) and SAHV-M2 (P<.05) (**Table 2**).

**Figure 3.**
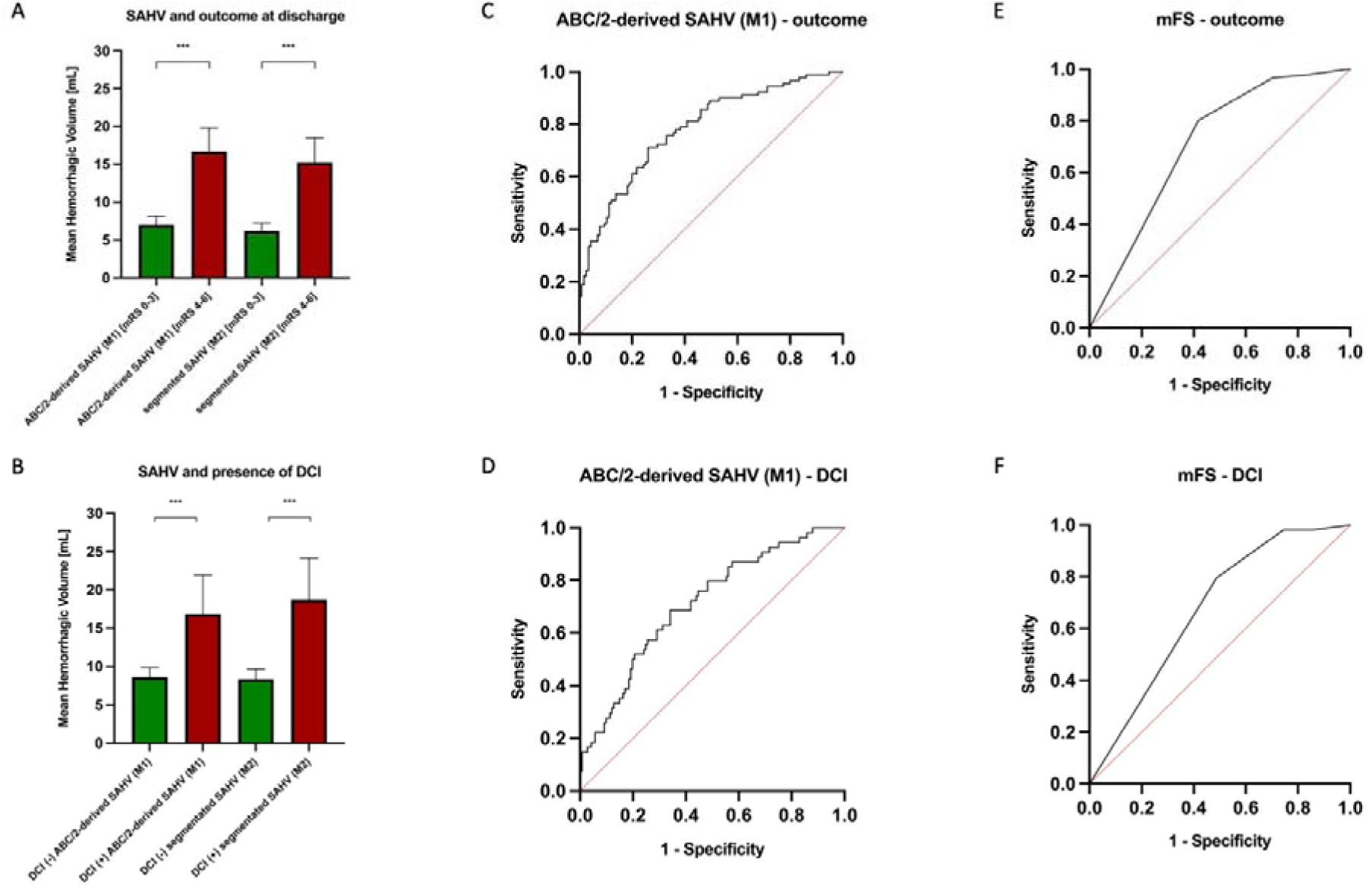
Relationship of Estimated Hemorrhagic Volumes With Target Variables. A and D, Mean volumes based on dichotomization into good outcome (mRS 0-3) and poor outcome (mRS 4-6) and the respective method (ABC-SAHV-M1= ABC/2-derived subarachnoid hemorrhage volume) applied to estimate these volumes. Results are shown in Table 2. B, ROC curve of ABC-SAHV-M1on outcome at discharge with AUC = 0.78 (95% CI, 0.71-0.84) C, ROC curve of mFS on outcome at discharge with AUC = 0.71 (95% CI, 0.64-0.78). E, ROC curve of ABC-SAHV-M1on DCI with AUC = 0.71 (95% CI, 0.63-0.79). F, ROC curve of mFS on presence of DCI with AUC = 0.67 (95% CI, 0.59-0.75). AUC indicates area under the curve; CHV, cisternal subarachnoid hemorrhage volume; DCI, delayed cerebral ischemia; mFS, modified Fisher Scale; mRS, modified Rankin scale; ROC, receiver operating characteristic.

**Table 2.** Differences in Mean Volumes By Functional Outcome and Presence of DCI mRS score.

|  | **ABC/2 derived -SAHV (M1)** | **segment-SAHV (M2)** |
| --- | --- | --- |
|  | **mean, mL** | **mean, mL** |
| mRS score |  |  |
| 0-3 | 7.01 (95% CI, 5.91-8.12) | 6.16 (95% CI, 5.10-7.22) |
| 4-6 | 16.63 (95% CI, 13.49-19.77) | 15.23 (95% CI, 11.98-18.48) |
| *P value*<sup>a</sup> | <.0001 | <.0001 |
| DCI |  |  |
| Yes | 8.66 (95% CI, 7.47-9.85) | 8.34 (95% CI, 7.06 -9.62) |
| No | 16.84 (95% CI, 11.74-21.94) | 18.23 (95% CI, 13.11-23.35) |
| *P value*<sup>a</sup> | <.0001 | <.0001 |
| Abbreviations: M1=model 1, ABC/2-derived-SubArachnoid Hemorrhage Volume (SAHV); M2= model 2, segment-SAHV = manually segmented SAHV; DCI, delayed cerebral ischemia; mRS, modified Rankin scale; |  |  |
| <sup>a</sup> Significance testing: unpaired *t* test |  |  |

ROC curve analysis (**Figure 3**) demonstrated that SAHV-M1 overall demonstrated a moderate to high discriminative and predictive strength for outcome, with an AUC of 0.78 (95% CI, 0.71-0.84). To account for the capabilities of these variables to predict DCI, logistic regression analysis and ROC curve analysis were performed separately. For this analytical segment, 10 patients had to be excluded due to death within 72 hours after admission. Therefore, 195 patients were included in this study analysis. ROC curve analysis of ABC/2-derived SAHV for DCI revealed an AUC of 0.71 (95% CI, 0.63-0.79) (**Figure 2 C and D**). These AUC data suggest that SAHV-M1 predicts outcomes and DCI more accurately than the modified Fisher scale (**Figure 3E and F**).

### Determination of SAHV Cutoff Values with Outcomes Using Youden Index

The optimum cut point is defined as the highest Youden index within a given set of ROC curve data and represents the maximal sensitivity and specificity cut point. Cutoff values were determined based on the predictor variable SAHV-M1 estimated for outcome and DCI. For both outcome and DCI, a cutoff value of 10 mL SAHV-M1 (J_ABC-SAHV-outcome_ = 10.0855 mL and J_ABC-SAHC-DCI_ = 10.117 mL) was determined in this study population (**Figure 4**). Results of the sensitivity, specificity, positive predictive and negative predictive for outcome and DCI using a cut-off value of 10 mL ABC/2-derived SAHV are shown in Table 3. This data suggest that a minimum “dose-response” relationship might exist of the amount of SAH blood within the basal cisterns with subsequent modified Rankin scale outcomes and development of DCI.

**Figure 4.**
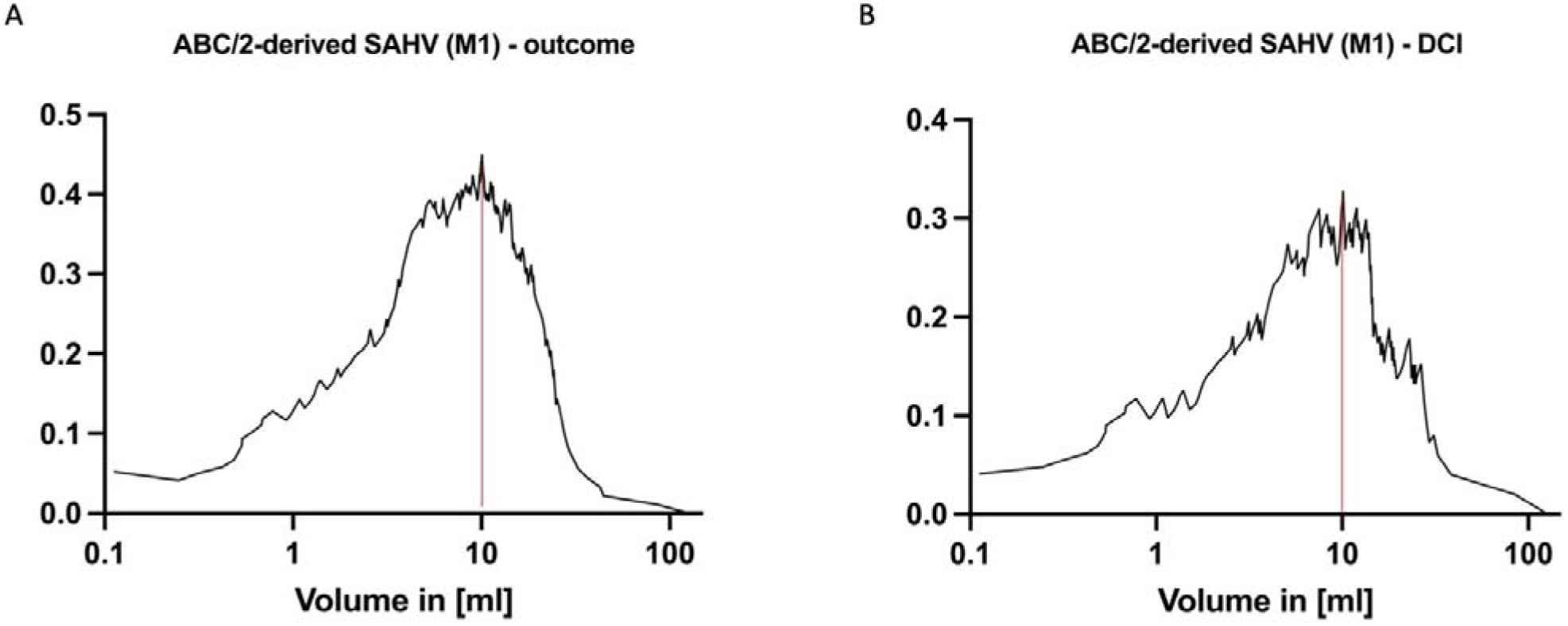
ABC-SAH-M1, Youdens Index (J = sensitivity + specificity – 1) for each value of predictor variables, dashed lines indicate maximum Youden’s index a.) J_ABC-SAHV_-outcome = 10.0855 mL; b.) J_ABC-SAHV_-DCI = 10.117

**Table 3.** Sensitivity, Specificity and Predictive Values of ABC2/derived SAHV (M1) cut-off volume (10 mL)

|  | **Outcome** | **DCI** |
| --- | --- | --- |
| *Sensitivity (%)* | 71.11 | 69.81 |
| *Specificity (%)* | 75.67 | 66.9 |
| *PPV (%)* | 69.56 | 40.5 |
| *NPV (%)* | 76.99 | 87.29 |

Abbreviations: DCI = Delayed cerebral ischemia; PPV = positive predictive value; NPV = negative predictive value

## Discussion

This study aimed to develop a simplified quantitative and more precise volumetric method to determine SAHV blood volume in aneurysmal SAH. We explored and compared two models and their association with outcomes and DCI. The SAH ABC/2-derived volumetric methodology appears similar to the well-established ABC/2 volumetric models for intracerebral hemorrhage which is powerfully predictive on outcomes [14, 15, 17]. Therefore, this SAH volumetric research has critical importance to advance future SAH translational research, and to our knowledge, is the first attempt to establish a clinically applicable, easy-to-use quantitative SAHV methodology similar to ABC/2 estimates for intracerebral hemorrhage. Further, prior studies have quantified SAH blood primarily through the application of automated or manual imaging segmentation software[12, 13, 19] which is not available in many emergency departments or widely utilized in clinical practice. We demonstrated that a simple ABC/2-derived SAHV method can predict poor outcomes by hospital discharge and a higher risk of future DCI during hospitalization. In a comparative AUC analysis, we found that the ABC/2-derived SAHV-M1 is at least equal or better in predicting outcome and DCI compared to the well-known modified Fisher scale which is prone to underestimate the true volumetric relationship of SAHV on clinical outcomes. Furthermore, comparative volumetric descriptions showed that the ABC/2-derived SAHV-M1 can approximate complex and time-consuming manual segmentation (SAHV-Model 2) on average. Although some differences between the estimated volumetrics of SAHV-M1 and other segmentation methods are evident, these discrepancies might be explained by difficult to define neuroanatomical SAH spaces on the gyri, sulci, and hemispheric convexities of the brain as well as a non-linear shaped ventricular system.[5, 19]

Lastly, we were able to identify a potential SAHV cutoff value using Youden’s index of 10 mL by SAHV-M1 which was associated with a higher risks of poor outcome and DCI. This preliminary SAHV cutoff value may be particularly useful for future larger, prospective, randomized-controlled SAH research trials and requires more validation. The identification of SAH volume as a continuous variable is clinically significant since it can be used to risk-stratify SAH patients and to establish a robust “dose-response” relationship for future neuroprotective therapies and using an individualized dosing approach. Furthermore, precise SAHV measurement helps provide a more robust and reliable baseline of blood volume and location to potentially perform targeted therapy with alternative routes of intrathecal or intraventricular drug delivery, or even adjunctive drug therapy [20]. The potential discovery of a minimal SAHV dose of 10mL associated with poor outcomes is clinically relevant and could become a target for future interventions and/or used as predictive tool for a quantifiable risk assessment of DCI.

The ABC/2-derived method of measuring intracerebral hemorrhage was found to be a powerful predictor on patient outcomes and became incorporated at comprehensive stroke centers[21] as a severity of illness in this disease. Similarly, we think that ABC/2 for measuring SAHV can be easily protocolized and standardized at both primary and comprehensive stroke centers. Our ABC/2-derived SAHV does not require any coding expertise, costly imaging software, nor machine-learning algorithms. A basic medical imaging view station of the NCCT with simple linear measurement tools is sufficient to determine the ABC’s of SAH blood in the 5 main “star-shaped” major cisterns. This simple method could be used by clinicians worldwide (eg, physicians, nurse practitioners, and radiologists) wherever the NCCT is available, and even in rural and underserved areas without comprehensive stroke centers.

Measurement and calculation of ABC/2-derived SAHV (Model 1) is similar to the learning curve for measuring ICH. In contrast, a manual segmentation approach takes much longer depending on the complexity and extent of SAH distribution. While manual segmentation has been demonstrated to be a quantifiable and a precise method in radiologic evaluation of SAH, [12] it remains highly demanding in terms of the overall time investment to manually segment slice by slice. Manual segmentation is not practical in most emergency SAH patient scenarios. Manual segmentation is useful mostly for comparative analysis in SAH research to help compare, derive, and develop primarily automated segmentation models.[22] Therefore, while not clinically applicable, manual segmentation approaches like SAH-M2 can still be used in prospective randomized trials to further validate our SAHV-M1 or other future SAHV models.

When comparing our ABC/2-derived model (M1) with established radiological frameworks such as the mFS and FS, one frequently expressed aspect of critique pertains to the substantial interobserver variability observed in the mFS when grading NCCT. This inherent variability subsequently undermines the robustness of its predictive validity. Recent data suggests that mFS achieves only moderate interobserver agreement with overall interrater reliability (IRR) by Kendell’s coefficient varying between Kw = 0.41 (95 % CI, 0.33 – 0.49) and Kw = 0.586 (p < 0.0005).[10, 23, 24] It is hypothesized that the IRR is negatively influenced by the vague definitions of mFS grading criteria as the hemorrhagic extent was described as either thin or thick. No specific metric characterization was proposed, thereby naturally compromising the model’s reproducibility.[7-9] While not evidently based, we again do think that a more quantitative approach, like the ABC/2-derived SAHV method, would allow for increased reproducibility. Additionally, both the mFS and original FS arguably lack discriminative accuracy between individual groups, therefore would not allow for reasonable quantifiable risk assessment, as it is our observation that the majority of patients diagnosed with aSAH are either graded as mFS 3 or mFS 4. This would notably restrict the potential of relevant therapeutic abstractions within in the realm of clinical decision making. However, given the inherent characteristics of the “dose-response” relationship existing between SAHV and functional outcomes or secondary complications [4, 6, 12, 13], we think that our ABC/2-derived model would represent a more comprehensive portrayal of this association. Consequently, it is plausible that our model would be better poised to support within the decision-making process regarding management of aSAH patients.

We acknowledge that our ABC/2-derived SAHV model 1 has limitations due to the complex morphology of SAH blood patterns and cisternal anatomy, which do not fully represent an ellipsoidal shape. Therefore, this mathematical simplification and volumetric estimation of SAH should be considered purely a volumetric approximation compared to more time-intensive segmentation methods that allow for almost pixel-level measurement beyond these 5 major cisterns. However, we feel this finding is important since it provides clinicians and researchers a relatively simple to use tool for SAH patients similar to ICH and subdural hematoma (SDH) that also use ABC/2 -derived formulas[14, 15, 17]. Prior literature has shown how three linear measurements (A, B, and C), even in crescent shaped SDH works as an approximation due to subtraction of two ellipsoid shapes to make one simplified shape.[14] There are certainly more complicated ways to measure these 5 major SAH neuroanatomic spaces and yet they likely still require splitting up each cisternal space into multiple (smaller) compartmentalized volumes and summation of these values. Lastly, volumetric estimation of SAH beyond the cisternal level was not performed using our ABC/2-derived SAH Model 1. Mathematically, this is because, it is our observation that as SAH blood approaches the cerebral convexities the actual volume becomes extremely small (i.e., Lim x -,’I 0) and contributes little to the main star-shaped cisternal SAH blood volume pattern seen on NCCT scans and what holds most quantifiable SAH blood. Also, the measurement of tiny distal neuroanatomic spaces would add tremendous methodical complexity and add considerable time costs, thereby decreasing potential clinical applicability and scalability.

It was not possible during this initial study to determine IRR for ABC/2-derived SAHV nor evaluate the methodological reproducibility of all the different mathematical systems of measurement. However, drawing upon the findings of prior investigations, it is probable that future studies could develop a more quantitative methodology and compare IRR among models, and compare older semiquantitative models such as the Fisher and modified Fisher scale.[23-25]

Lastly, we were not able to analyze IPH, SDH and IVH-associated SAH blood patterns separately but we plan to incorporate these hemorrhage types into a future larger study. Of note, the number of large visible IVH and SDH patterns were much less frequent in our data set and would not provide a statistically significant power or sample size for our primary SAH study. Therefore this would limit our ability to abstract any clinically relevant or predictive capabilities on outcomes due to the small sample size of these cases. Also, we note a recent study showed that IPH did not significantly impact a model’s predictive value when analyzed independently[12].

## Conclusion

A simplified ABC/2-derived SAH volumetric (SAHV) model using admission non-contrast head CT estimates blood volume similar to segmentation methods and is predictive of discharge modified Rankin scale outcomes and future risk of DCI. A potential SAHV cutoff value of 10 mL or greater of SAH blood appears to predict higher risk of poor clinical course and DCI. SAH blood volume (SAHV) should be validated similar to the ICH score by Hemphill et al but for SAH patients. Similarly, we believe an ABC/2-derived model of SAH blood measurement could spark future translational SAH research, more robust SAH research in neuroprotective drugs and interventions, and help more precisely quantify whether SAH blood volume has a dose-response relationship on future clinical outcomes and DCI.

## Data Availability

All data produced in the present study are available upon reasonable request to the authors

## Acknowledgments

The authors thank Alison Dowdell in the section of Scientific Publications at Mayo Clinic who helped provide copyediting, proofreading, administrative, and academic support.

## Supplemental Tables

**Table 1a:** Logistic regression analysis of ABC/2- derived SAHV (M1)

|  | Univariate logistic regression analysis |  |  |  |  | Adjusted logistic regression analysis** |  |
| --- | --- | --- | --- | --- | --- | --- | --- |
|  | Odds Ratio<br>(CI 95 %) | Wald | Significance | R<sup>2</sup>* | AUC | Odds Ratio<br>(CI 95 %) | Sig. |
| **Poor outcome** | 1.158<br>(1.107-1.218) | 35.572 | <0.001 | 0.307 | 0.779<br>(0.715-0.842) | 1.143<br>(1.087-1.201) | < 0.001 |
| **DCI** | 1.094<br>(1.049-1.140) | 17.932 | < 0.001 | 0.159 | 0.709<br>(0.630-0.789) | 1.105<br>(1.052-1.160) | < 0.001 |

Abbreviations: DCI = Delayed cerebral ischemia
\*Pseudo-R-squared measured via Nagelkerke R square
\*\* EXP(B) in multivariable analysis adjusted for gender, age, smoking, alcohol abuse, hypertension, hyperlipidemia, diabetes mellitus, CVD, COPD;

