## Supplementary material for "The ABCs of Subarachnoid Hemorrhage Blood Volume Measurement: A Simplified Quantitative Method Predicts Outcomes and Delayed Cerebral Ischemia": Table 1a

|  |  | Modified Fisher Scale/ original Fisher Scale | ABC/2 derived -SAHV (M1) | | Manually segmented-SAHV (M2) |
| --- | --- | --- | --- | --- | --- |
| Positive Characteristics |  | - Low time investment - Does not require any specialized medical imaging software | | - Does not require any specialized medical imaging software - Allows for a quantifiable risk assessment of outcome and DCI - Likely higher IRR due to quantitative methodology | - Most precise method of volumetric SAHV measurement - The entire extent of SAHV can be measured - High IRR - Potential to be integrated into AI based models |
| Negative Characteristics |  | - Descriptive Grouping - Poor IRR - Questionable discriminative accuracy between groups - Tends to underestimate the true volumetric relationship - Little to no consequence in clinical decision making | | - Moderate time investment - Cannot account for the extension of SAHV beyond the cisternal level - Simplified volumetric approximation | - High time investment - Requires specialized medical imaging software and expertise |

Supplementary Files

**Table 1a** Outline of positive and negative aspects of different radiologic models
